# Prenatal phthalate mixture exposure increases early childhood internalising problems via maternal oxidative stress

**DOI:** 10.1101/2025.11.09.25339835

**Authors:** Sarah Thomson, Katherine Drummond, Martin O’Hely, Thomas Boissiere-O’Neill, Christos Symeonides, David Burgner, Toby Mansell, Richard Saffery, Peter J. Vuillermin, Peter D. Sly, the BIS Investigator Group, Anne-Louise Ponsonby

**Affiliations:** The Florey Institute of Neuroscience and Mental Health, Parkville, Victoria, Australia; The University of Melbourne, The Florey Department of Neuroscience and Mental Health, Parkville, Victoria, Australia; Deakin University, Institute for Mental and Physical Health and Clinical Translation (IMPACT), Geelong, Victoria, Australia; Murdoch Children’s Research Institute, Parkville, Victoria, Australia; The University of Queensland, Child Health Research Centre, Brisbane, Queensland, Australia; The University of Melbourne, Department of Paediatrics, Parkville, Victoria, Australia; Monash University, Department of Paediatrics, Clayton, Victoria, Australia; Barwon Health, Child Health Research Unit, Geelong, Victoria, Australia

**Keywords:** Prenatal, phthalate, environmental mixture, oxidative stress, child mental health, mediation, birth cohort

## Abstract

Prenatal phthalate exposure has been linked to internalising problems in children. Maternal oxidative stress is a plausible biological mechanism underlying this association as it is induced by phthalates and is associated with early internalising problems, but this pathway has not yet been empirically tested. In the Barwon Infant Study, a population-based birth cohort of 1,074 Australian children, we investigated the potential mixture effect of prenatal phthalate exposure on maternal oxidative stress and assessed whether oxidative stress mediates the relationship between the prenatal phthalate mixture and early internalising problems. Concentrations of phthalate metabolites and nucleic acid oxidation biomarkers were measured in third-trimester maternal urine, and phthalate daily intakes estimated. Internalising problems were assessed at ages 2 and 4 years using the parent-report Child Behavior Checklist and Strengths and Difficulties Questionnaire, respectively. Applying weighted quantile sum regression with repeated holdout validation, we found that higher phthalate mixture exposure was associated with increased maternal oxidative stress (0.22 standard deviations per interquartile range increase in mixture, 95% CI: 0.13, 0.31), with dimethyl phthalate and diethyl phthalate as main contributors. Bayesian kernel machine regression yielded comparable results. For all child outcomes, counterfactual mediation analyses across holdout datasets revealed evidence of mediating effects, suggesting that oxidative stress is a pathway through which prenatal phthalate mixture exposure increases early-childhood internalising problems. These findings highlight the heightened risk associated with co-exposure to multiple phthalates during pregnancy and the potential to mitigate this harm not only through improved chemical regulation but also by monitoring and reducing maternal oxidative stress.

**Graphical abstract:** 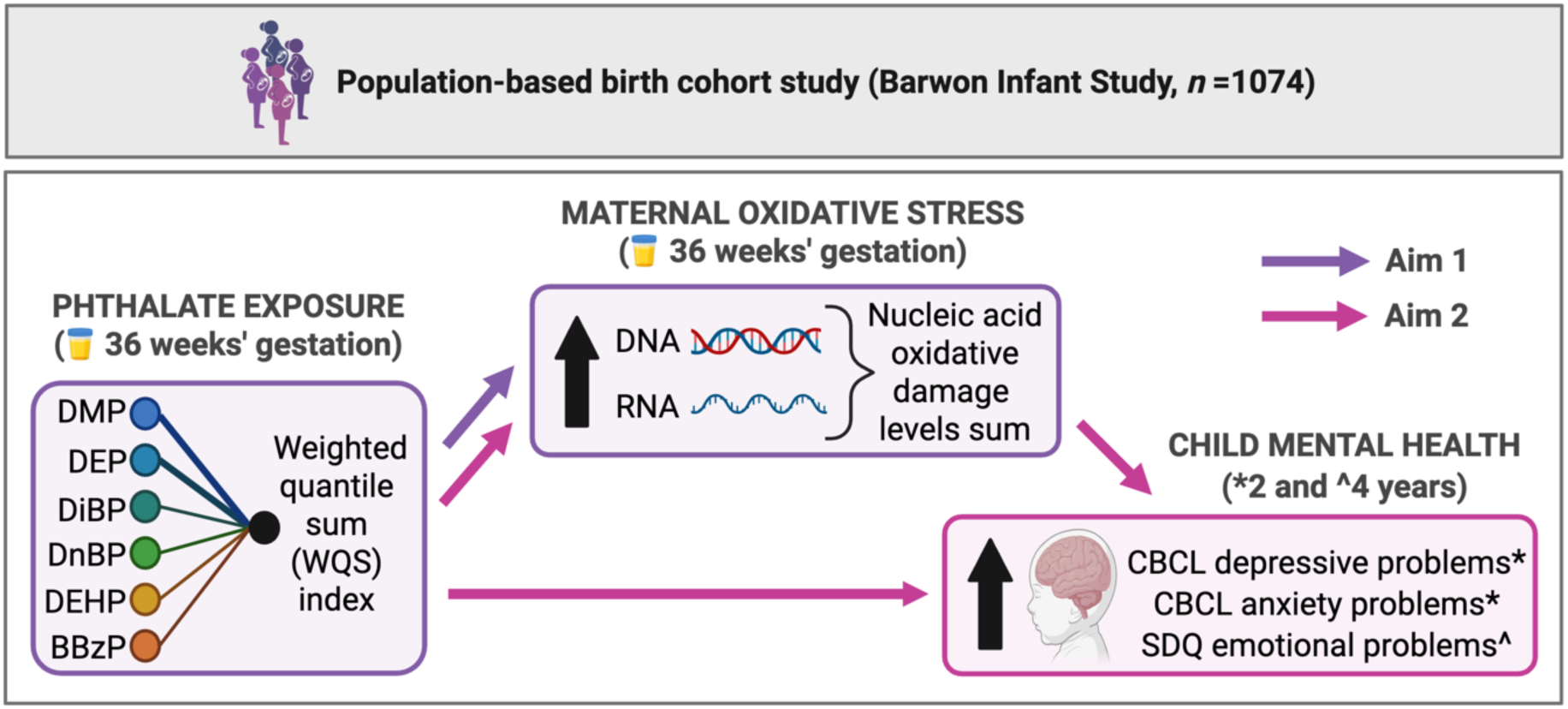

## 1. Introduction

Mental disorders are the leading cause of disability in young people.^1^ Anxiety and depressive disorders are among the most common, affecting 6.9% and 2.8% of 4- to 17-year-olds in Australia, respectively.^2^ Internalising behaviour, in which distress and negative emotions are directed inwards, is a core feature of anxiety and depression and may develop early in life, with signs evident from one year of age.^3^ Children who display early internalising problems are at increased risk of anxiety and depressive disorders in childhood,^4^ adolescence,^4^ and adulthood.^5^ The prenatal period is critical for the development of brain structures and neural circuits underlying emotional regulation.^6^ During this time, the rapid growth and plasticity of the foetal brain render it particularly susceptible to environmental insults, such as exposure to toxicants.^6^

Phthalates are a class of synthetic, industrial chemicals widely used across consumer products.^7^ They are ubiquitous in the environment, and their metabolites are detectable in the urine of nearly all individuals.^8,9^ Phthalates of low molecular weight (LMW), such as dimethyl phthalate (DMP), diethyl phthalate (DEP), di-isobutyl phthalate (DiBP) and di-n-butyl phthalate (DnBP), primarily function as solvents, stabilisers, fixatives and film-forming agents in cosmetics, personal care products, pharmaceuticals, adhesives and paints.^7^ High-molecular-weight (HMW) phthalates, such as di(2-ethylhexyl) phthalate (DEHP) and butyl benzyl phthalate (BBzP), are primarily used as plasticisers in polyvinyl chloride-based products including food packaging, medical devices, vinyl flooring, cables and building materials.^7^ As phthalates are not chemically bound to the products in which they are incorporated, they can leach and off-gas into the surrounding environment.^10^ Human exposure to LMW phthalates is predominantly through inhalation and dermal absorption, whereas HMW phthalates are primarily ingested.^11^

Epidemiological studies have repeatedly linked higher prenatal urinary phthalate metabolite levels with internalising problems in early childhood,^12–18^ although null findings have also been reported.^19–21^ Investigating potential underlying biological mechanisms represents an important next step.^22^ Oxidative stress, mitochondrial dysfunction, and inflammation have emerged as key interconnected pathways of interest in relation to prenatal phthalate exposure and mental disorders more broadly.^23–26^ Here, we focus on oxidative stress, building on our prior findings that maternal urinary biomarkers of oxidative stress are associated with early childhood emotional problems.^27^

Oxidative stress occurs when the production of reactive oxygen species (ROS) exceeds the capacity of antioxidant and/or repair systems, resulting in cellular damage.^28^ Experimental studies show that certain phthalates can increase ROS production and impair antioxidant defences.^29–34^ In epidemiological research, oxidative stress is commonly assessed using stable urinary biomarkers, such as 8-hydroxy-2ʹ-deoxyguanosine (8-OHdG) and 8-hydroxyguanosine (8-OHG) which reflect oxidative damage to DNA and RNA, respectively.^35–37^ Numerous studies have reported positive associations between individual phthalates and oxidative stress biomarkers.^38^ Positive associations have also typically been reported between phthalate mixtures and oxidative stress biomarkers in pregnant^39–41^ and non-pregnant populations,^42,43^ but data are limited. A ‘mixture’ refers to a group of environmentally co-occurring exposures that may act together additively or interactively to affect a biological process or health outcome. Given that phthalates are ubiquitous and individuals are exposed to multiple phthalates simultaneously, it is essential to study mixture effects.

Several studies have employed causal mediation analysis to demonstrate that oxidative stress mediates the relationship between exposure to individual phthalate metabolites and adverse health outcomes across multiple systems, including cardiometabolic, endocrine, reproductive, and gastrointestinal domains.^44–47^ These findings strengthen causal arguments and suggest that oxidative stress is a common mechanistic pathway by which phthalates exert harmful effects on the human body. A limited number of studies have focused on prenatal exposure and adverse child outcomes, with mediating effects being reported for pre-term birth^48^ and impaired cognitive development.^49^ To date, no study has investigated oxidative stress as a mediator between prenatal exposure to phthalates—either individually or as a mixture—and child internalising problems. More broadly, research on molecular pathways mediating the effects of environmental chemical mixtures on human health remains limited, in part due to a lack of suitable statistical methods. Yet advancing this area is critical because shared biological pathways may represent targets for secondary prevention.^50^ This is particularly important given the ubiquity of phthalates and other environmental chemicals and the challenges of achieving primary prevention.

Therefore, our aims in this study were to: (i) investigate the potential association between phthalate mixture exposure and oxidative stress during pregnancy using weighted quantile sum (WQS) regression with repeated holdouts, and (ii) evaluate whether, and to what extent, maternal oxidative stress mediates the effect of this phthalate mixture on early childhood internalising problems, with total, indirect and direct effects estimated using a counterfactual mediation framework embedded in the repeated holdout WQS regression from aim (i).

## 2. Methods

### 2.1 Study design and participants

The Barwon Infant Study (BIS) is an ongoing prospective study following parents and children from pregnancy to school age. The BIS design, recruitment and population characteristics have been described previously.^51^ Briefly, 1,074 mother–child pairs (10 sets of twins) were recruited at or before 28 weeks of gestation from June 2010 to June 2013 using an unselected antenatal sampling frame in the Barwon region of Victoria, Australia. Women were excluded if they were under the age of 18 years, not Australian permanent residents, or required an interpreter to complete questionnaires.^51^ Infant exclusion criteria included birth before 32 weeks’ gestation, serious illness within the first few days of life, major congenital malformations, or genetically determined disease.^51^ The study was approved by the Barwon Health Human Research Ethics Committee (HREC 10/24) and families provided written informed consent. For the current analysis, the second-born twin in each twin set (n=10) was excluded, leaving 1,064 mother–child pairs.

### 2.2 Maternal phthalate metabolite measurement

Methods of collection, analysis and processing have been previously described.^52,53^ Briefly, a single spot urine specimen was collected from participants at 36 weeks of gestation and collaborators at the Queensland Alliance for Environmental Health Science used high-performance liquid chromatography/tandem mass spectroscopy to quantify urinary phthalate metabolite levels. We restricted analysis to phthalate metabolites detected above the limit of detection (LOD) in at least 90% of the cohort. For these metabolites, measurements below the LOD (0–7%) were imputed with the LOD divided by the square root of 2.^54^ To correct for possible batch effects, the fractional deviations between batch-specific and overall geometric means of quality control measures were used to adjust phthalate measurements.^55^ To account for urine dilution, specific gravity was measured and the Levine–Fahy equation applied.^56^ The time of day of sample collection was accounted for by regressing the phthalate concentrations on time of day and retaining the residuals.^57^ From the processed urine concentrations, phthalate estimated daily intakes (EDIs) were calculated accounting for estimated maternal weight at urine collection, fractional excretion of the compound, and compound-to-metabolite molecular weight ratio.^53^ An EDI for DMP was calculated from the urinary metabolite monomethyl phthalate (MMP); DEP from monoethyl phthalate (MEP); DiBP from mono-isobutyl phthalate (MiBP); DnBP from mono-n-butyl phthalate (MnBP); BBzP from monobenzyl phthalate (MBzP); and DEHP from the metabolites mono(2-ethyl-5-hydroxyhexyl) phthalate (MEHHP), mono(2-ethyl-5-oxohexyl) phthalate (MEOHP), and mono(2-ethyl-5-carboxypentyl) phthalate (MECPP). Given that the distribution of each phthalate EDI was approximately log-normal, a log_2_ transformation was applied to normalise the data for analysis.

### 2.3 Maternal oxidative stress biomarkers

Collaborators at the Australian National Phenome Centre (Perth, WA, Australia) used liquid chromatography–mass spectrometry (LC–MS/MS) to quantify levels of 8-OHdG and 8-OHG in maternal urine collected at 36 weeks of gestation. Detailed methods have been reported previously.^27^ Briefly, an ExionLCTM system (SCIEX; Framingham, MA, USA), a Kinetex C8 2.6μm 2.1×150mm column (Phenomenex; Lane Cove West, NSW, Australia), and a QTRAP 6500+ system (SCIEX; Framingham, MA, USA) were used for chromatographic separation, reversed-phase separation at 40°C, and mass spectrometry detection with electrospray ionisation, respectively. Analyst®1.7.1 was used for data acquisition and SCIEX OS Analytics 1.7.0 software (SCIEX; Framingham, MA, USA) for analysis. Measurements below the limit of quantification (LOQ) were imputed with the LOQ divided by the square root of 2.^54^ Raw urinary 8-OHdG and 8-OHG urinary concentrations were corrected for: (i) batch by applying weights based on the fractional deviation of batch-specific from overall geometric mean quality control values,^55^ (ii) urine dilution using specific gravity measurements and the Levine–Fahy equation,^56^ and (iii) the time interval between urine collection and storage by regressing the oxidative stress marker concentrations on the time interval and retaining the residuals.^57^ The oxidative stress variables each approximated a log-normal distribution and were log_2_ transformed for analysis.

### 2.4 Child internalising problems

#### 2.4.1 Child Behavior Checklist at age two years

The Child Behavior Checklist (CBCL) for Ages 1.5 to 5 years is a widely used questionnaire to assess emotional and behavioural problems in pre-school aged children.^58^ It consists of 99 descriptive statements, and parents must rate how well each item describes their child now or within the past 2 months using a three-point Likert scale (0 = not true; 1 = somewhat or sometimes true; 2 = very true or often true). The CBCL has good psychometric properties with evidence of validity as well as high test-retest reliability, inter-rater reliability, and internal consistency.^58,59^ It includes seven syndrome subscales, three composite scales, and five Diagnostic and Statistical Manual of Mental Disorders, Fifth Edition (DSM-5)-oriented subscales, which together are used to evaluate children’s behaviour. Here, we focused on the DSM-5-oriented depressive subscale and the DSM-5-oriented anxiety subscale which are predictive of anxiety and depressive disorders.^60^ As recommended for regression analyses, the raw scores, instead of the scaled T-scores, were used.^58^

#### 2.4.2 Strengths and Difficulties Questionnaire at age four years

The Strengths and Difficulties Questionnaire (SDQ) P4–10 is a commonly used, 25-item questionnaire completed by parents to assess emotional and behavioural problems in children aged four to ten years.^61^ Each item is a descriptive statement to which parents respond based on the child’s behaviour over the past six months using a three-point Likert scale (0 = not true; 1 = somewhat true; 2 = certainly true). In a meta-analysis of 48 studies, the SDQ demonstrated strong psychometric properties.^62^ Five subscale scores and three summary scale scores are used to evaluate behaviour. As the SDQ does not have separate anxiety and depression scales, we focused on the emotional problems subscale, which is moderately to strongly predictive of anxiety and depressive disorders.^63^

### 2.5 Statistical analysis

All statistical analyses were conducted using R version 4.4.1. Where relevant, statistical significance was defined as a *p*-value below 0.05.

#### 2.5.1 Oxidative stress composite score

To generate a composite score reflecting nucleic acid oxidation (RNA and DNA damage) as a proxy for oxidative stress, we standardised the 8-OHG and 8-OHdG variables, summed them, and re-standardised the resulting composite. The Pearson correlation between 8-OHG and 8-OHdG was 0.64 and the composite score captured 82% of the total variance in the two markers. It was used as a proxy for oxidative stress in all subsequent analyses and is referred to as ‘oxidative stress’.

#### 2.5.2 Weighted quantile sum regression: Phthalate mixture effect on oxidative stress

To evaluate the association between exposure to a phthalate mixture and maternal oxidative stress in pregnancy, and to identify phthalate chemicals of concern, we used WQS regression with repeated holdouts. WQS regression is a method for analysing environmental mixtures that constructs a weighted linear combination of exposure variables (the WQS index) and evaluates its association with an outcome in a regression model.^64^ In more detail, exposures are categorised using quantiles, and the data are randomly partitioned into training and holdout subsets. In the training data, exposure weights and the index regression coefficient are estimated by constrained non-linear optimisation with weights restricted to be non-negative and sum to one, and the mixture effect constrained to a pre-specified direction. To stabilise weight estimates, this optimisation is repeated across bootstrap samples. The final WQS index is then applied in the holdout data, where the outcome is regressed on the index to obtain the mixture–outcome association. To reduce overfitting, this procedure can be repeated across multiple training–holdout splits.^65^

In our implementation, we used deciles to categorise the phthalates and linear regression to model oxidative stress. We hypothesised a positive mixture effect *a priori* based on previous research.^39–43^ We set the number of bootstrap samples to 100 and repeated the WQS procedure across 100 random splits of the data (25% training, 75% holdout). We report mean beta and weight estimates with normal approximation 95% confidence intervals (CIs) calculated using the standard deviation of estimates across holdout sets. We also report percentile-based estimates and CIs (median, 2.5^th^, 97.5^th^ percentiles) to account for potential departures from normality. Chemicals of concern were identified using criteria from Busgang et al.: a chemical was deemed a *probable* contributor if its weight exceeded the equal-weight threshold (1/*c* where *c* is the number of chemicals) in >90% of holdout sets, and a *possible* contributor if this occurred in >50–90%.^66^ Analyses were conducted using the *gWQS* R package.^67^

#### 2.5.3 Mediation analysis: Indirect effect of phthalate mixture on internalising problems via oxidative stress

To investigate whether prenatal phthalate mixture exposure affects child internalising problems in early childhood via maternal oxidative stress in pregnancy, we conducted counterfactual mediation analysis using the phthalate mixture indices from our repeated holdout WQS regression of oxidative stress. Counterfactual mediation analysis decomposes the total effect of an exposure on an outcome into the portion that operates through an intermediate variable—the natural indirect effect—and the portion that operates independently—the natural direct effect.^22^ It assumes no unmeasured confounding of the exposure–mediator, mediator–outcome and exposure–outcome relationships, and no exposure-induced mediator–outcome confounding. The final WQS index in repeated holdout WQS regression is constructed using weights averaged across all training sets, thereby incorporating information from the full dataset. Using this index in a mediation analysis on the same sample risks overfitting and biased inference. To address this, we integrated mediation analysis into the repeated holdout procedure by conducting it in each of the 100 holdout sets using the WQS index calculated from weights estimated in the corresponding training set. As with WQS regression, we report mean estimates across the 100 holdout sets as well as percentile-based estimates. See **Figure 1** for method infographic. Mediation analyses were conducted using the *medflex* R package.^68^ A linear regression model was used for the mediator, and a quasi-Poisson regression model was used for the outcome, given that both the CBCL and SDQ raw scores are summed ordinal measures that approximate counts of behavioural symptoms and exhibit right-skewed, overdispersed distributions.

**Figure 1.**
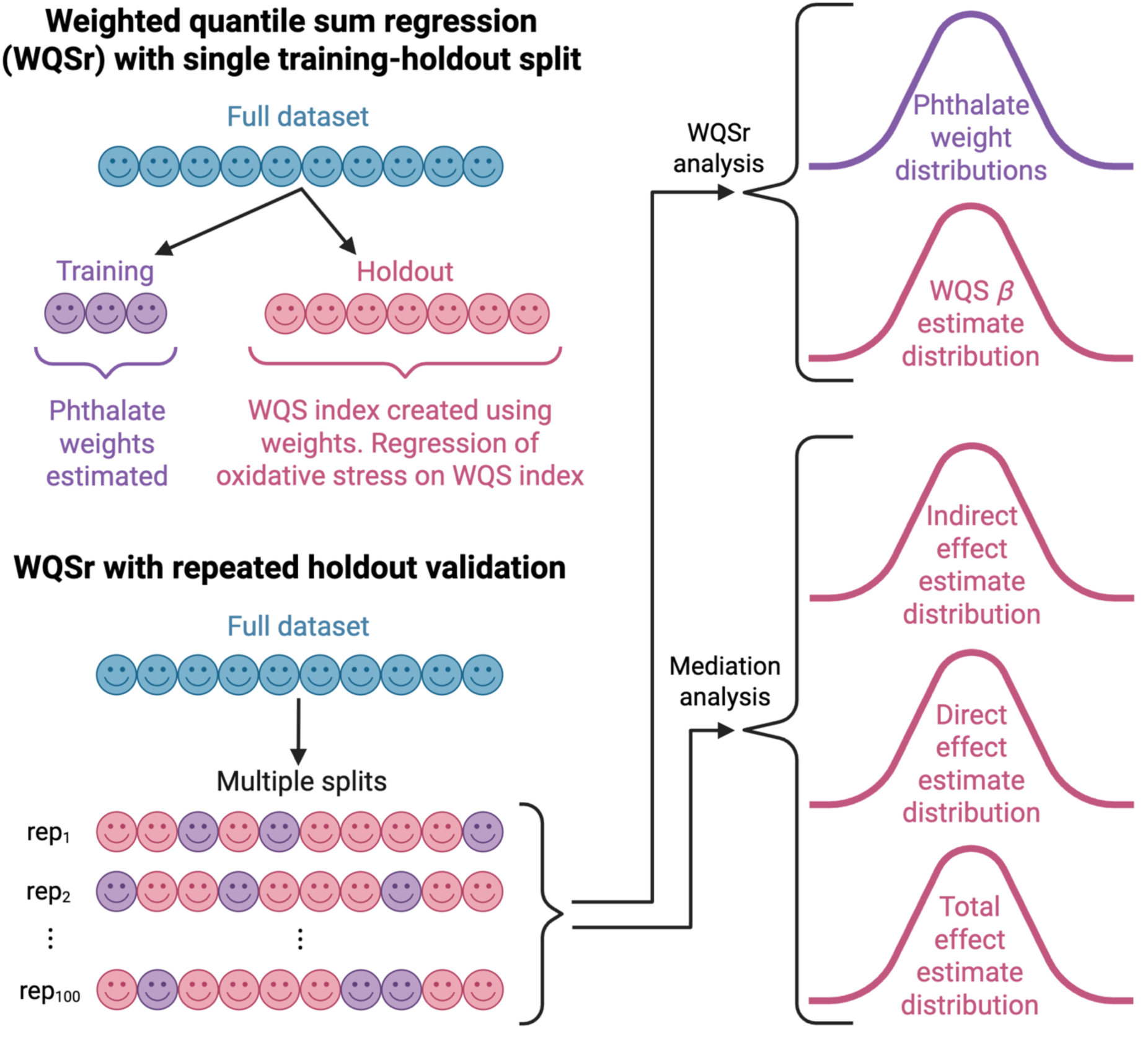
Infographic illustrating the repeated holdout weighted quantile sum (WQS) regression approach and the integration of causal mediation analysis (CMA) in the present study. The data were repeatedly split into training and holdout subsets. Weights for each phthalate were estimated in the training sets and applied to the corresponding holdout sets to calculate the weighted sum (WQS index) and to estimate its association with oxidative stress. This process yielded distributions of phthalate weight and WQS coefficient estimates, from which mean estimates were obtained. CMA was then performed in each holdout set using the WQS index (calculated from weights estimated in the corresponding training set) as the exposure, oxidative stress as the mediator, and an internalising measure as the outcome. This process yielded distributions of indirect, direct, and total effect estimates, from which mean estimates were obtained.

#### 2.5.4 Sensitivity analysis: Bayesian kernel machine regression to assess phthalate mixture effect on oxidative stress

To assess the robustness of results to violations of the linearity and additivity assumptions in the WQS regression model, we employed a second, more flexible model: Bayesian kernel machine regression (BKMR).^69^ BKMR estimates the relationship between an exposure mixture and an outcome with an exposure–response function governed by a Gaussian process prior. The covariance structure of this prior is defined by a Gaussian kernel, which reflects similarities in individuals’ exposure profiles. Together these allow the exposure–response function to be estimated as a smooth but flexible surface, accommodating complex non-linearities and interactions. Prior distributions are also placed on the kernel parameters, covariate regression coefficients, and residual variance.

Markov chain Monte Carlo (MCMC) sampling combines priors with the observed data, yielding a posterior distribution for the exposure–response function. In our application of BKMR, variable selection was enabled, and default diffuse priors were used for the global variance scale, variable inclusion probability and residual variance. To guide the choice of prior for the smoothing parameters (*r_m_*), we fitted separate kernel machine regression models for each exposure, fixing *r* at different values and inspecting the resulting smoothness. We ran four MCMC chains, each with 60,000 iterations and a burn-in of 30,000 iterations. The Metropolis–Hastings tuning parameters were adjusted to achieve an acceptance rate of 20–50%. The *bkmr*, *bkmrhat* and *coda* R packages were used to implement BKMR and run MCMC diagnostics.^70–72^

#### 2.5.5 Additional analyses and other sensitivity analyses

In additional analyses, we tested how each phthalate daily intake associates with oxidative stress and the child internalising problem measures in separate regression models. As some participants had missing data (**Table S1**) and the primary analyses were based on available cases, a sensitivity analysis was conducted using multiple imputation under the assumption that data were missing at random rather than missing completely at random. We implemented multiple imputation by chained equations (MICE) with the *mice* R package,^73^ imputing missing values for all participants (n=1064) and generating 100 imputed datasets. The imputation model included all variables from the inferential analyses to preserve their associations, as well as auxiliary predictors of the incomplete variables and of missingness (**Table S1**).^74^ To investigate if mediation effects differed by child sex, we ran sex-stratified analyses. While maternal genetics were not available in this cohort, we have previously generated a child pathway-based genetic score for oxidative stress.^26^ We assessed the informativeness of this score for the mediation pathway by testing its association with maternal oxidative stress and whether it modified the association between maternal oxidative stress and child internalising problems.

#### 2.5.6 Model covariates

All models included the following covariates: gross annual household income in pregnancy (<$75,000 AUD vs. ≥$75,000), maternal age (years), reported ethnicity (all four grandparents Caucasian vs. not), genetic ancestry (first principal component [PC1] from a principal component analysis [PCA] of genome-wide SNP genotyping data), parity (primiparous vs. multiparous), pre-pregnancy BMI (kg/m^2^), active smoking (any in pregnancy vs. none), passive smoking (daily exposure in any trimester—T1, T2 or T3—vs. otherwise), alcohol use (≥1 standard drinks in T1, T2 and T3 vs. otherwise), maternal diet (PC1 from PCA of 99 food frequency questionnaire items completed at 28 weeks’ gestation),^75^ gestational age at urine collection (weeks), and child’s sex assigned at birth.

Models involving the child internalising outcomes additionally included the following covariates: child’s age at behavioural assessment (years) and maternal perceived stress (Perceived Stress Scale, PSS–14, score at 28 weeks’ gestation). Maternal pregnancy conditions (preeclampsia, hypertension, and gestational diabetes mellitus) were not adjusted for due to low prevalence (<5% of the sample); however, participants with these conditions were excluded in a sensitivity analysis.

## 3. Results

### 3.1 Participant flow and sample characteristics

A participant flowchart for BIS is presented in **Figure S1**. Summary characteristics of the inception cohort as well as the sample used in the mixture analysis in this paper are displayed in **Table 1**. In the cohort, the mean age of the mothers at conception was 31 years. Approximately three quarters of the children were reported as having four grandparents of European descent, and around one quarter had two university-educated parents. High proportions (93%–100%) of the mothers whose urine was tested for phthalates in the inception cohort had detectable levels of the phthalate metabolites. Among the mothers tested for oxidative stress markers, 93% and 86% had quantifiable levels of 8-OHG and 8-OHdG, respectively.

**Table 1.** Participant characteristics.

|  | Cohort sample<br>(N=1064) |  | Mixture analysis sample<br>(N=634) |  |
| --- | --- | --- | --- | --- |
|  | N | Mean (SD)<br>or % (n)<br>or Median [IQR]<br>or GM {GSD} | N | Mean (SD)<br>or % (n)<br>or Median [IQR]<br>or GM {GSD} |
| **Constitutional and socio-economic factors** |  |  |  |  |
| Mother's age at conception (years) | 1,064 | 31.3 (4.8) | 634 | 31.9 (4.4) |
| Annual household income under \$75K AUD at 28w gestation | 1,031 | 30.0% (309) | 634 | 23.3% (148) |
| Mother and father university-educated | 1,050 | 26.5% (278) | 632 | 31.5% (199) |
| SEIFA index of relative socio-economic disadvantage at 28w | 1,051 | 1029 [988-1064] | 627 | 1031 [996-1071] |
| Single-parent household at 28w gestation | 1,061 | 4.1% (43) | 634 | 2.1% (13) |
| All four grandparents of European descent | 1,050 | 72.8% (764) | 634 | 75.6% (479) |
| Multiparous | 1,064 | 55.3% (588) | 634 | 55.4% (351) |
| Child's sex assigned at birth male<sup>1</sup> | 1,064 | 51.8% (551) | 634 | 53.9% (342) |
| **Prenatal factors** |  |  |  |  |
| Pre-pregnancy BMI (kg/m<sup>2</sup>) | 917 | 25.4 (5.5) | 634 | 25.0 (5.2) |
| Any active smoking in pregnancy | 948 | 11.3% (107) | 607 | 6.9% (42) |
| Any passive smoking in pregnancy | 904 | 15.3% (138) | 585 | 11.1% (65) |
| Alcohol consumed in all trimesters | 1,041 | 12.9% (134) | 627 | 15.5% (97) |
| Gestational diabetes mellitus | 900 | 4.9% (44) | 528 | 3.6% (19) |
| Pre-eclampsia | 1,029 | 3.3% (34) | 616 | 2.1% (13) |
| Gestational hypertension | 1,022 | 2.6% (27) | 617 | 2.3% (14) |
| Perceived Stress Scale score at 28w gestation | 801 | 18.7 (7.0) | 549 | 17.7 (6.7) |
| Gestational age at urine collection (weeks) | 928 | 36.3 (0.7) | 634 | 36.2 (0.7) |
| **Plastic chemical exposure at 36w (µg/kg body weight/day)** |  |  |  |  |
| DMP daily intake | 842 | 0.0 {2.4} | 634 | 0.0 {2.4} |
| DEP daily intake | 842 | 1.6 {3.8} | 634 | 1.6 {3.7} |
| DiBP daily intake | 842 | 0.8 {2.1} | 634 | 0.8 {2.1} |
| DnBP daily intake | 842 | 1.0 {2.1} | 634 | 1.0 {2.0} |
| DEHP daily intake | 842 | 1.6 {2.1} | 634 | 1.6 {2.1} |
| BBzP daily intake | 842 | 0.2 {3.0} | 634 | 0.2 {2.9} |
| **Oxidative stress urinary levels at 36w (ng/mL)** |  |  |  |  |
| 8-OHG | 812 | 18.4 {2.9} | 634 | 18.8 {2.9} |
| 8-OHdG | 809 | 3.6 {2.2} | 634 | 3.7 {2.2} |
| **Birth factors** |  |  |  |  |
| Gestational age at birth (weeks) | 1,064 | 39.5 (1.5) | 634 | 39.7 (1.2) |
| Caesarean birth | 1,064 | 30.6% (326) | 634 | 29.8% (189) |
| Child birthweight (kg) | 1,062 | 3.5 (0.5) | 634 | 3.6 (0.5) |
| **Child affective problems** |  |  |  |  |
| CBCL/1.5-5 DSM-5-oriented depressive subscale (raw score) | 671 | 1 [0-2] | 486 | 1 [0-2] |
| CBCL/1.5-5 DSM-5-oriented anxiety subscale (raw score) | 671 | 1 [0-3] | 486 | 1 [0-3] |
| Child's age at CBCL/1.5-5 assessment (years) | 672 | 2.5 (0.1) | 486 | 2.4 (0.1) |
| SDQ/P4-10 emotional problems subscale | 787 | 1 [0-2] | 542 | 1 [0-2] |
| Child's age at SDQ/P4-10 assessment (years) | 787 | 4.1 [4.0-4.2] | 542 | 4.1 [4.0-4.2] |

NB. SD, standard deviation; IQR, interquartile range; GM, geometric mean; GSD, geometric standard deviation; w, weeks; SEIFA, Socio-Economic Indexes for Areas; BMI, body mass index; DMP, dimethyl phthalate; DEP, Diethyl phthalate; DiBP, Diisobutyl phthalate; DnBP, Di-n-butyl phthalate; DEHP, Di-(2-ethylhexyl) phthalate; BBzP, butyl benzyl phthalate; 8-OHG, 8-hydroxyguanosine; 8-OHdG, 8-hydroxy-2'-deoxyguanosine; CBCL, Child Behavior Checklist; DSM, Diagnostic and Statistical Manual of Mental Disorders; SDQ, Strengths and Difficulties Questionnaire. <sup>1</sup> There were no known intersex children in the cohort.

### 3.2 Weighted quantile sum regression: Phthalate mixture effect on oxidative stress

Using WQS regression, an association between the phthalate mixture and maternal oxidative stress was detected (**Figure 2**, **Table S2**). Specifically, mean maternal oxidative stress levels were estimated to increase by 0.22 standard deviation (SD) units (95% CI: 0.13, 0.31) per interquartile range (IQR) increase in the phthalate WQS index, holding all other variables in the model constant. **Figure 3** and **Table S3** summarise the distribution of phthalate weights across the 100 repeated holdouts. DMP made the largest contribution to the mixture with a mean weight of 0.43 and 92% of its estimated weights above the equal weight threshold, making it a *probable* contributor according to criteria from Busgang et al.^66^ The next largest was DEP (mean weight=0.20) which had weight estimates above the equal weight threshold 57% of the time and was therefore a *possible* contributor. For each of the other phthalate chemicals, more than 50% of the estimated weights were below the equal weight threshold.

**Figure 2.**
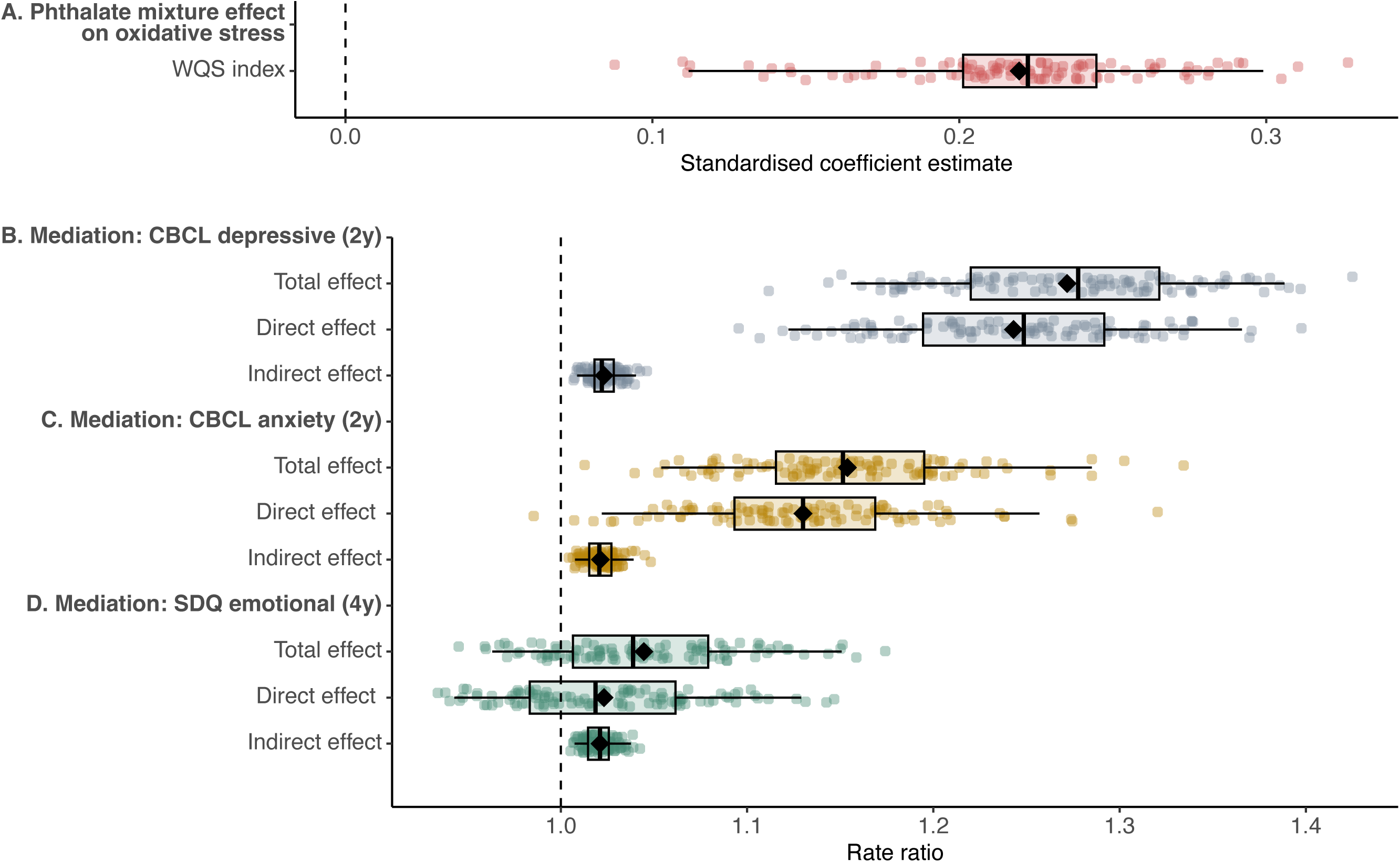
Estimated phthalate mixture effect on maternal oxidative stress in pregnancy from weighted quantile sum regression analysis with 100 repeated holdouts (**A**). Causal mediation analysis estimates in 100 holdouts, with corresponding WQS index from analysis A as exposure, oxidative stress as mediator, and child CBCL DSM-5-oriented depressive problems subscale score at 2 years as outcome (**B**) or child CBCL DSM-5-oriented anxiety problems subscale score at 2 years as outcome (**C**) or child SDQ emotional problems subscale score at 4 years as outcome (**D**). Data points show the standardised estimates in each of the 100 holdouts. Box plots summarise the distribution of the estimates across the 100 holdouts (box represents the 25th, 50th, and 75th percentiles, and whiskers extend to the 2.5th and 97.5th percentiles). Closed diamonds represent the mean estimates for the 100 holdouts.

**Figure 3.**
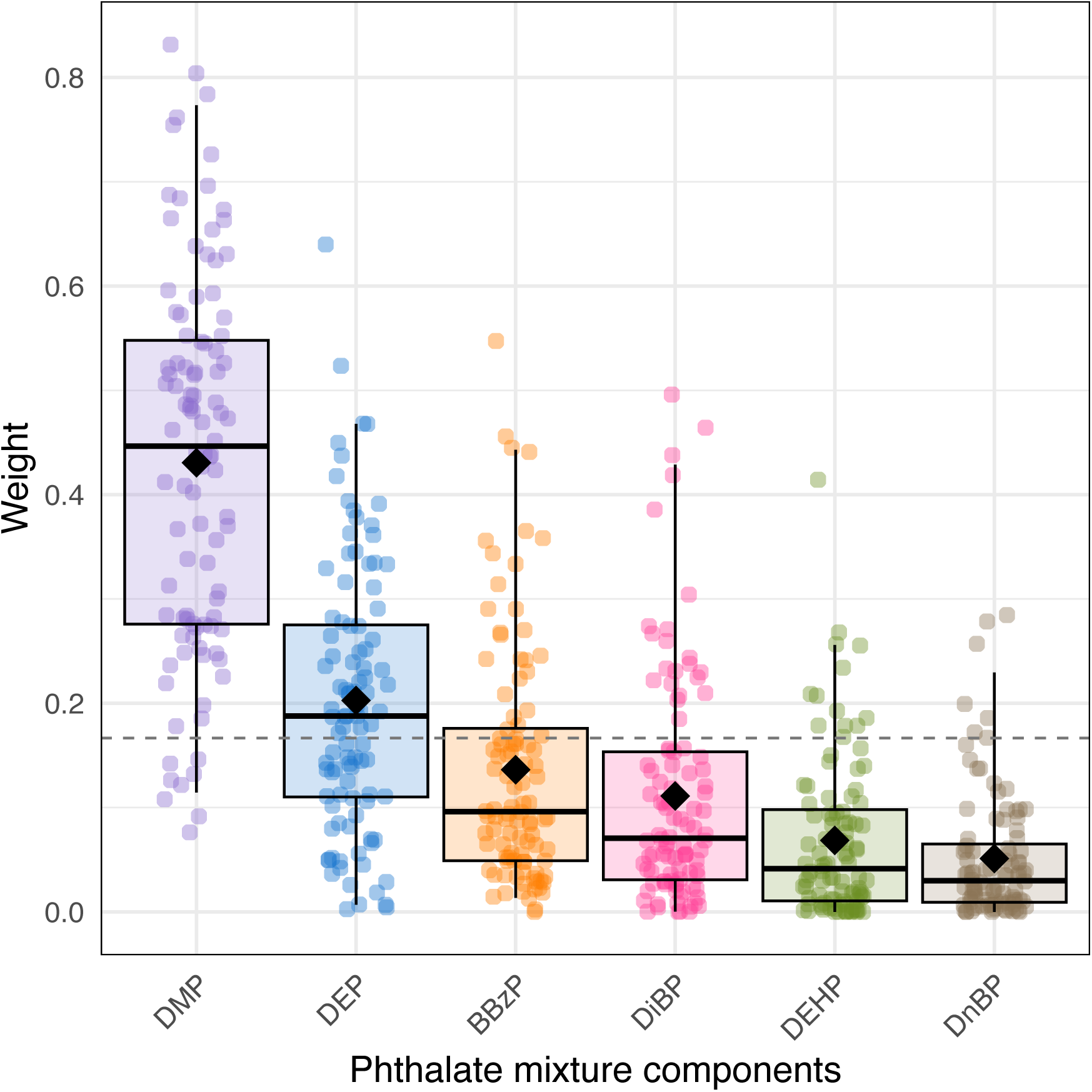
Identification of phthalate chemicals of concern in relation to maternal oxidative stress in pregnancy using weighted quantile sum regression with repeated holdouts. Data points show the weights in each of the 100 holdouts. Box plots summarise the distribution of the weights across the 100 holdouts (box represents the 25th, 50th, and 75th percentiles, and whiskers extend to the 2.5th and 97.5th percentiles). Closed diamonds represent the mean weights for the 100 holdouts. The dotted line shows the expected weight (1/6=0.17) if all six phthalate compounds contributed equally to the mixture. *Abbreviations:* DMP, dimethyl phthalate; DEP, diethyl phthalate; BBzP, butyl benzyl phthalate; DiBP, diisobutyl phthalate; DEHP, di-(2-ethylhexyl) phthalate; DnBP, di-n-butyl phthalate.

### 3.3 Mediation analyses: Indirect effect of phthalate mixture on internalising problems via oxidative stress

Pooling the total effect estimates from the mediation analyses across the holdout sets, the expected CBCL depressive problems score and CBCL anxiety problems score were estimated to increase by 27% (IRR=1.27, 95% CI: 1.14, 1.41) and 15% (IRR=1.15, 95% CI: 1.04, 1.27), respectively, per IQR increase in the phthalate WQS index. The respective estimated proportions of these effects mediated by oxidative stress were 10% (indirect effect: incidence rate ratio IRR=1.02, 95% CI: 1.01, 1.04) and 15% (indirect effect: IRR=1.02, 95% CI: 1.00, 1.04). The pooled total effect estimate for the SDQ emotional problems subscale measured at 4 years was smaller in magnitude (IRR=1.04, 95% CI: 0.95, 1.15) but the indirect effect, representing the pathway from the prenatal phthalate mixture to child emotional problems via maternal oxidative stress in pregnancy, was similar (IRR=1.02, 95% CI: 1.01, 1.04) giving an the estimated proportion mediated of 49%. Plots and summaries of the distributions of the mediation estimates across the 100 holdout sets are presented in **Figure 2** and **Table S2**. To test normality assumptions, we also calculated percentile-based estimates and CIs (median, 2.5th, 97.5th percentile) which were almost identical to the original estimates in the WQS regression and mediation analysis (**Table S2–S3**).

### 3.4 Sensitivity analysis: Bayesian kernel machine regression to assess phthalate mixture effect on oxidative stress

The Bayesian kernel machine regression analysis, used to assess the robustness of results to violations of the underlying assumptions of the WQS regression model, also showed a mixture effect (**Figure 4A**). Specifically, a change from the 25th percentile to the 75th percentile in all phthalate daily intake variables was associated with a 0.21 (95% credible interval: 0.02, 0.39) SD increase in oxidative stress. Posterior inclusion probabilities, which provide a measure of exposure importance, were highest for DMP, followed by DEP, consistent with the WQS regression findings (**Table S4**).

**Figure 4.**
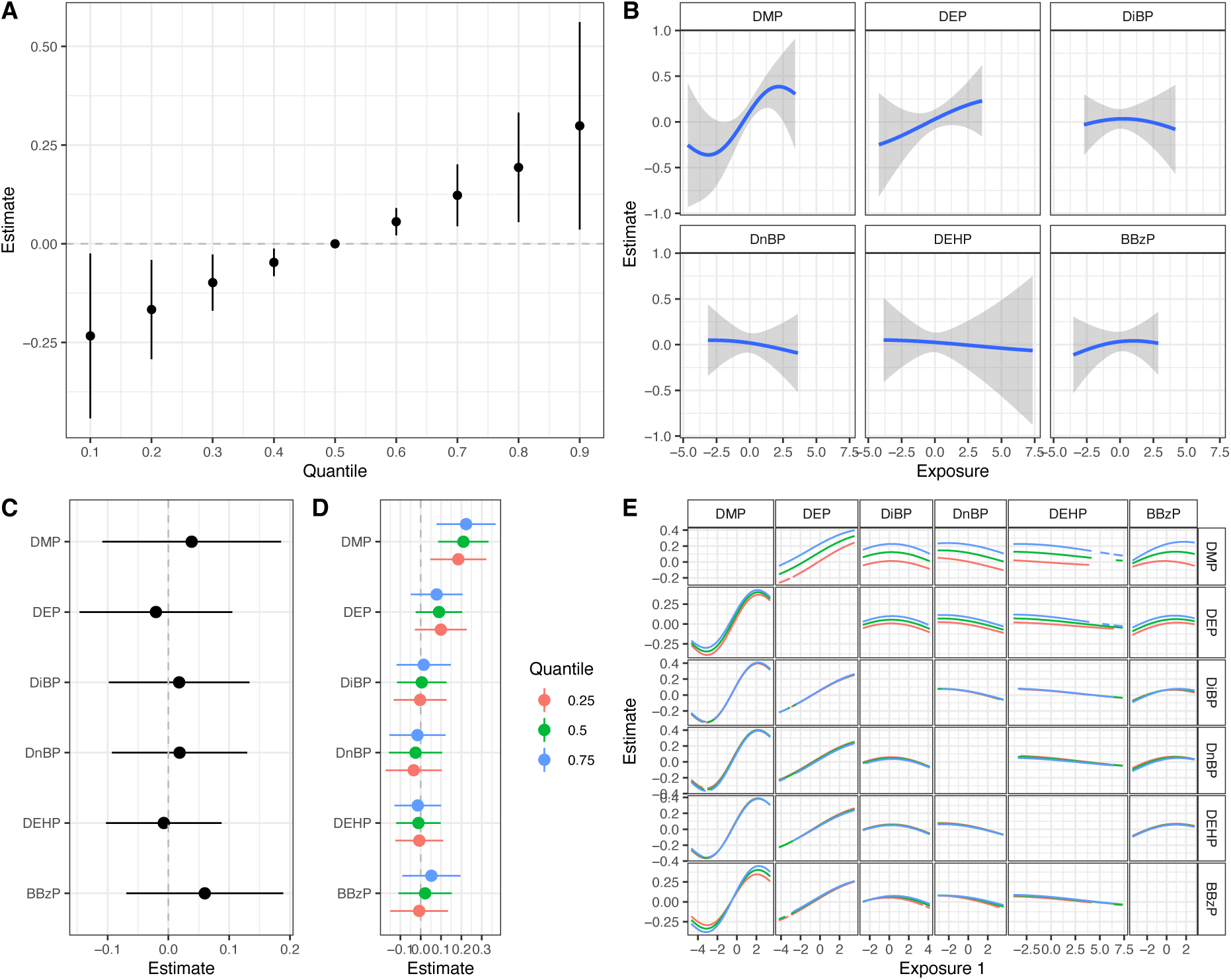
Bayesian kernel machine regression results. **A.** The estimated change (with 95% credible interval) in oxidative stress for a change in all phthalate exposures from their median values to the percentile showing on the x-axis. **B.** Univariate exposure-response function (shaded area indicating 95% credible interval) for each phthalate exposure and oxidative stress while all other phthalate exposures are fixed at their median value. **C.** The estimated change (with 95% credible interval) in oxidative stress for a change in a phthalate exposure from its 25^th^ to the 75^th^ percentile while all other phthalate exposures are fixed at the absolute difference between their 75^th^ and 25^th^ percentiles. **D.** The estimated change (with 95% credible interval) in oxidative stress for a change in a phthalate exposure from its 25^th^ to the 75^th^ percentile while all other phthalate exposures are fixed at their 25^th^, 50^th^ or 75^th^ percentiles. **E.** Exposure-response function for a phthalate exposure and oxidative stress when a second phthalate exposure is at its 25^th^, 50^th^ or 75^th^ percentile and all other phthalate exposures are fixed at their median value. *Abbreviations:* DMP, dimethyl phthalate; DEP, diethyl phthalate; DiBP, diisobutyl phthalate; DnBP, di-n-butyl phthalate; DEHP, di-(2-ethylhexyl) phthalate; BBzP, butyl benzyl phthalate.

Exposure–response functions for single phthalate exposures, with the other phthalates fixed at their median values, showed linear or near-linear relationships with oxidative stress (**Figure 4B**). Some curvature was evident for DMP at the ends of the distribution; however, a linear pattern was encompassed by the 95% credible interval (**Figure 4B**). The BKMR analysis provided little evidence of interaction between the phthalates (**Figure 4C–E**). For each phthalate, an IQR increase was associated with similar changes in oxidative stress, regardless of whether the other phthalate exposures were fixed at their (i) 25th, (ii) 50th or (iii) 75th percentiles (**Figure 4D**). **Figure 4C** plots the difference in the scenario (i) and (iii) estimates for each phthalate, with the largest differences observed for DMP and BBzP. However, all 95% credible intervals crossed the line of no effect. When stronger evidence of interaction is observed, a plot of bivariate exposure–response functions can help identify which pairs of exposures are interacting (**Figure 4E**). For all pairs, the exposure– response functions for the first phthalate and oxidative stress—when the second phthalate was fixed at its (i) 25th, (ii) 50th, and (iii) 75th percentiles, and all others were fixed at their medians—were equidistant or near-equidistant suggesting no interaction (**Figure 4E**).

### 3.5 Additional analyses and other sensitivity analyses

As an additional analysis, we regressed oxidative stress and the three child internalising problem measures on each of the phthalates separately. The results are presented in **Table S5** and all estimates are for an IQR change in the phthalate. Consistent with the mixture results, the two top-ranked phthalates associated with oxidative stress, based on effect size and *p*-value, were DMP (β=0.14 SD, 95% CI: 0.06, 0.22) and DEP (β=0.13 SD, 95% CI: 0.03, 0.23). DMP was also associated with the CBCL depressive score (IRR=1.19, 95% CI: 1.07, 1.30) and DEHP with the CBCL anxiety score (IRR=1.09, 95% CI: 1.00, 1.18). For the phthalates and SDQ emotional scale, none of the tests reached statistical significance.

To address the potential for selection bias in our results, we imputed missing data using multiple imputation and re-ran the WQS regression and mediation analysis. A similar pattern of results was observed. The pooled estimates across the holdouts and imputed datasets are presented in **Table S6** and **Table S7**. **Figure S2** illustrates the distribution of mean WQS regression and mediation estimates (averaged across holdouts) across the imputed datasets. **Figure S3** shows the same for the mean WQS component weight estimates. We also assessed whether mediation effects differed by child sex using sex-stratified analyses. Estimates were similar for boys and girls, with slightly larger point estimates observed for boys in the CBCL outcomes (**Table S8**). However, the confidence intervals overlapped substantially, suggesting that there were no meaningful sex-specific differences. While maternal genetic data was not available, we assessed whether a previously derived child oxidative stress pathway genetic score was informative for the mediation pathway. It was not associated with maternal oxidative stress (β=0.22, 95% CI –0.07, 0.51) and did not modify the association between maternal oxidative stress and child internalising problems across the three outcomes (interaction IRRs 0.97–1.01, all *p*>0.80). In a final sensitivity analysis, the WQS regression and mediation models were repeated after excluding mothers with pre-eclampsia, hypertension, and gestational diabetes mellitus. The findings were essentially unchanged (results not shown).

## 4. Discussion

In a population-based birth cohort, we found that higher prenatal phthalate mixture exposure was associated with increased maternal oxidative stress at 36 weeks of gestation. Low-molecular-weight (LMW) phthalates contributed most to the mixture, particularly DMP, with a smaller contribution from DEP. This phthalate mixture was also associated with child internalising problems at age 2 years, but there was no evidence of such at age 4. However, at both time points, we observed a positive indirect effect through maternal oxidative stress providing the first evidence of greater prenatal phthalate mixture exposure increasing child internalising problems at least in part by elevating maternal oxidative stress in pregnancy.

Our findings are consistent with experimental evidence that individual phthalates can induce oxidative stress. *In vitro* studies demonstrate that DEHP, DEP, and DnBP increase ROS production in various human cell types, including hepatic (HepG2), endometrial stromal, and placental cells,^29,30,32^ and that DEHP can decrease expression of key antioxidant enzymes.^29^ *In vivo* studies further support these effects: across multiple tissues, oral DEHP and DnBP exposures in rodents increase ROS, reduce antioxidant enzyme activity, and induce lipid and DNA oxidative damage.^31,33,34^ Mechanistic pathways supported in the literature include disruption of mitochondrial function and the electron transport chain, leading to excess mitochondrial ROS;^76^ peroxisome proliferator-activated receptor (PPAR) activation, which increases peroxisomal β-oxidation and generates hydrogen peroxide as a by-product;^77^ cytochrome P450–mediated oxidative metabolism of HMW phthalates, producing ROS via uncoupling of the CYP catalytic cycle;^78,79^ epigenetic suppression of antioxidant defence genes, reducing ROS-neutralising capacity;^80^ and activation of inflammatory signalling pathways, promoting cytokine release and ROS production via immune cell oxidases.^81^ However, oxidative stress, mitochondrial dysfunction, and inflammation are interconnected processes that engage in dynamic feedback loops, making it difficult to distinguish between initiating events and reinforcing effects.^24^

Our findings are also consistent with several epidemiological studies reporting positive associations between individual phthalates and oxidative stress biomarkers in both pregnant ^82–85^ and non-pregnant populations.^38^ Among these biomarkers, 8-OHdG has been frequently examined and is consistently associated with several phthalate compounds.^38,84,85^ Despite the potential for additive and interactive effects of phthalates, relatively few studies have assessed phthalate mixtures in relation to oxidative stress, reporting consistent, positive associations with 8-OHdG,^39,41,43^ and both positive and null associations for lipid peroxidation markers;^39–43^ however, small sample sizes were a limitation (n=105–156).^41,43^ In our analysis, LMW phthalates DMP and DEP were the primary contributors to the phthalate mixture. This aligns with previous studies in which LMW phthalates often drove the mixture effects, although DEHP metabolites were sometimes implicated.^40–43^ We also observed linear or near-linear exposure–response relationships, consistent with findings by Zeng et al., who reported linear associations between third-trimester, log-transformed phthalate and 8-OHdG levels in single-phthalate models.^41^ Our study extends these findings by demonstrating similar patterns within a mixture framework using BKMR, which accounts for co-exposure of multiple phthalates.

Previous research investigating a link between maternal oxidative stress and child internalising problems is very limited. In BIS, we have previously reported positive associations of maternal nucleic acid oxidative damage markers with CBCL depressive symptoms at 2 years and SDQ emotional problems at 4 years.^27^ Here, we extend this to include anxiety symptoms at 2 years. In contrast, Rommel et al. found no evidence of an association between third-trimester urinary lipid peroxidation markers and internalising problems at ages 4–5 as measured by the parent-rated Behavior Assessment System for Children (BASC–2).^86^ Discrepancies between studies may reflect differences in outcome measurement as our measures relate mainly to depression and anxiety whereas the BASC–2 internalising problems scale also captures social withdrawal. They could also reflect differences in oxidative stress measurement given that weak correlations have been reported among oxidative stress biomarkers with different molecular targets and clearance kinetics.^82,87^

There are several mechanisms by which maternal oxidative stress may influence offspring emotional behaviour. Since ROS have a short half-life, direct placental transfer of ROS from the mother is unlikely.^88^ However, elevated ROS levels can activate inflammatory cascades and the hypothalamic-pituitary-adrenal axis, which may affect the foetus directly through partial placental transfer of pro-inflammatory cytokines and cortisol,^89,90^ and indirectly via placental signalling.^91^ In addition, maternal oxidative stress can contribute to endothelial dysfunction, potentially impairing uteroplacental blood flow and restricting the delivery of nutrients and oxygen to the foetus.^92^ In the third trimester, a critical period for brain development, these pathways can induce foetal hypoxia, neuroinflammation and oxidative stress (to which the foetal brain is particularly susceptible due to immature antioxidant defences).^91,93^ They may also disrupt HPA axis programming, neurotransmitter systems, and both structural and functional development in regions involved in emotional regulation, such as the amygdala.^91,94^

Many epidemiological studies have reported positive associations between prenatal phthalate exposure and internalising problems in early childhood.^12–18^ We report similar positive associations, specifically for DMP and DEHP. Fewer studies have considered phthalate mixture effects reporting both significant^12,95^ and null findings^19,96,97^ for depression, anxiety or the internalising construct.

However, our phthalate mixture was defined based on its association with maternal oxidative stress, a biologically plausible mediator, rather than being optimised for prediction of child outcomes. This approach prioritises mechanistic relevance and reflects the temporal pathway from exposure to outcome. Despite not being outcome-optimised, the mixture still showed overall total effects for the CBCL subscales. The corresponding direct effect estimates suggest that other biological pathways may contribute at the 2-year timepoint but diminish in importance by age 4. In contrast, the comparable indirect effects across both ages imply that the oxidative stress pathway may exert a more enduring influence, although this requires further investigation. Differences in questionnaire instruments across the two time points may also have contributed to the observed discrepancies.

Current regulatory approaches to phthalates in the European Union (EU), the United States, Canada, and Australia restrict certain compounds, such as DEHP, BBzP, DiBP and DnBP, in children’s toys, cosmetics and, in the EU, a broader range of consumer products.^98–102^ In contrast, DMP and DEP remain largely unregulated, with restrictions applying only in Australia and limited to certain personal care products.^102^ These phthalates have historically been deemed safe at standard exposure levels, based largely on single-chemical assessments or cumulative risk evaluations focused on a narrow set of reproductive and developmental toxicity outcomes.^103^ However, in 2021, the European Chemicals Agency identified DMP and DEP as needing further regulatory consideration and also suggested a group-based regulatory approach to phthalates.^104^ Our findings add to a growing body of evidence indicating that exposure to DMP, DEP and phthalate mixtures in general may adversely affect human health in ways not adequately addressed by current regulatory frameworks.^41,43,85^

This study has several strengths. The assessment of a phthalate mixture instead of analysing individual compounds better reflects real-life exposure scenarios. By examining and finding statistical support for maternal oxidative stress as a molecular mechanism, the study strengthens the causal argument linking prenatal phthalate exposure to early childhood internalising problems. The integration of causal mediation analysis within the repeated holdout WQS regression framework avoided overfitting bias in the mediation estimates. Additionally, a flexible mixture method, BKMR, was employed to assess the robustness of our results to violations of the linearity and additivity assumptions in the WQS regression model. The consistency of results between the available-case analysis and the multiple imputation analysis reduces concerns about sampling bias, and comprehensive questionnaire data allowed for a rigorous assessment of potential confounders. To reduce potential measurement bias, we assessed and corrected for process factors related to the measurement of urinary markers. In terms of the child outcomes, the study utilised two widely validated and clinically relevant instruments—the CBCL and SDQ—to assess internalising problems in early childhood.

Our study also has some limitations. Only single spot urine samples were available. However, repeated measurements have shown phthalates to be moderately consistent over the third trimester;^105,106^ oxidative stress markers 8-OHdG and 8-OHG exhibit no significant diurnal variation and modest day-to-day variation;^36,107^ and random within-person variability would bias effect estimates toward the null. Only two biomarkers were used to create a composite oxidative stress measure; future studies could incorporate a broader panel for a more comprehensive assessment of systemic oxidative stress. Maternal genetic data were unavailable in this cohort, precluding incorporation of maternal genetic susceptibility to oxidative stress into our analyses. However, exploratory checks indicated that child oxidative stress pathway genetic score was not a suitable proxy for maternal susceptibility and did not modify the maternal oxidative stress and child internalising association. While child genetic modifiers could plausibly operate on the natural direct—and therefore total—effect, evaluating such modifiers was beyond the scope of this study. Six phthalate compounds were available in our cohort to evaluate as a mixture; however, other phthalates and other chemical classes—such as, bisphenols, parabens, pesticides, and per- and polyfluoroalkyl substances—may also have pro-oxidant effects,^108^ so we could not rule out confounding by these co-exposures or consider a more comprehensive chemical mixture effect. Future research should consider including a broader range of environmental chemicals in mixture analyses.

## 5. Conclusions

A positive association between exposure to a phthalate mixture and maternal oxidative stress in late pregnancy was observed, with LMW phthalates DMP and DEP contributing most strongly to the mixture effect. Causal mediation analysis suggests that greater exposure to this phthalate mixture increases child internalising problems at ages 2 and 4 years by elevating maternal oxidative stress.

These findings strengthen the causal evidence linking prenatal phthalate exposure to adverse foetal neurodevelopment and later mental health outcomes and suggest that third-trimester maternal oxidative stress may represent a modifiable target for secondary prevention. Future studies should aim to replicate and extend these findings by examining broader environmental mixtures that may act through oxidative stress pathways. Our novel integration of causal mediation analysis into the repeated holdout WQS regression framework offers a useful approach for investigating mediation in the context of environmental mixtures.

## CRediT authorship contribution statement

**Sarah Thomson**: Conceptualisation, Methodology, Sotware, Formal analysis, Data curation, Writing – original draft, Writing – review & editing, Visualization; **Katherine Drummond**: Conceptualisation, Writing – review & editing; **Martin O’Hely**: Methodology, Writing – review & editing; **Thomas Boissiere-O’Neill**: Writing – review & editing; **Christos Symeonides**: Conceptualisation, Writing – review & editing, Funding acquisition; **David Burgner**: Conceptualisation, Writing – review & editing; **Toby Mansell**: Writing – review & editing; **Richard Saffery**: Conceptualisation, Writing – review & editing; **Peter Vuillermin**: Conceptualisation, Resources, Writing – review & editing, Supervision, Project administration, Funding acquisition; **Peter Sly**: Conceptualisation, Writing – review & editing, Funding acquisition; **Anne-Louise Ponsonby**: Conceptualisation, Resources, Writing – original draft, Writing – review & editing, Supervision, Project administration, Funding acquisition.

## Data Availability

Barwon Infant Study (BIS) data requests are considered on scientific and ethical grounds by the BIS Steering Committee. If approved, data are provided under collaborative research agreements.

## Acknowledgements

The authors thank the BIS participants for the generous contribution they have made to this project. We also thank current and past staff for their efforts in recruiting and maintaining the cohort and in obtaining and processing the data and biospecimens. We acknowledge Barwon Health, Murdoch Children’s Research Institute, and Deakin University for their support in the development of this research. We also acknowledge the other members of the BIS Investigator Group including Lawrence Gray, Len Harrison, and Sarath Ranganathan, as well as past members John Carlin, Amy Loughman, Fiona Collier, Terry Dwyer, and Katie Allen for their contributions to BIS. Finally, we thank Chris Gennings for her generous feedback on a presentation of early results. Graphical abstract and Figure 1 were created with BioRender.com.

## References

1 Kieling, C. et al. Worldwide Prevalence and Disability From Mental Disorders Across Childhood and Adolescence: Evidence From the Global Burden of Disease Study. JAMA Psychiatry 81, 347–356 (2024). 10.1001/jamapsychiatry.2023.5051

2 Lawrence, D., et al. The mental health of children and adolescents. Report on the second Australian child and adolescent survey of mental health and wellbeing. (2015). <https://www.health.gov.au/sites/default/files/documents/2020/11/the-mental-health-of-children-and-adolescents_0.pdf>.

3 Briggs-Gowan, M. J., Carter, A. S., Bosson-Heenan, J., Guyer, A. E. & Horwitz, S. M. Are infant-toddler social-emotional and behavioral problems transient? J Am Acad Child Adolesc Psychiatry 45, 849–858 (2006). 10.1097/01.chi.0000220849.48650.59

4 Mesman, J. & Koot, H. M. Early preschool predictors of preadolescent internalizing and externalizing DSM-IV diagnoses. J Am Acad Child Adolesc Psychiatry 40, 1029–1036 (2001). 10.1097/00004583-200109000-00011

5 Reef, J., van Meurs, I., Verhulst, F. C. & van der Ende, J. Children’s problems predict adults’ DSM-IV disorders across 24 years. J Am Acad Child Adolesc Psychiatry 49, 1117–1124 (2010). 10.1016/j.jaac.2010.08.002

6 Tien, J., Lewis, G. D. & Liu, J. Prenatal risk factors for internalizing and externalizing problems in childhood. World J Pediatr 16, 341–355 (2020). 10.1007/s12519-019-00319-2

7 Schettler, T. Human exposure to phthalates via consumer products. Int J Androl 29, 134–139; discussion 181-135 (2006). 10.1111/j.1365-2605.2005.00567.x

8 Shin, H. M. et al. Temporal Trends of Exposure to Phthalates and Phthalate Alternatives in California Pregnant Women during 2007-2013: Comparison with Other Populations. Environ Sci Technol 54, 13157–13166 (2020). 10.1021/acs.est.0c03857

9 Sugeng, E. J. et al. Predictors with regard to ingestion, inhalation and dermal absorption of estimated phthalate daily intakes in pregnant women: The Barwon infant study. Environ Int 139, 105700 (2020). 10.1016/j.envint.2020.105700

10 Hahladakis, J. N., Velis, C. A., Weber, R., Iacovidou, E. & Purnell, P. An overview of chemical additives present in plastics: Migration, release, fate and environmental impact during their use, disposal and recycling. J Hazard Mater 344, 179–199 (2018). 10.1016/j.jhazmat.2017.10.014

11 Koch, H. M. et al. Identifying sources of phthalate exposure with human biomonitoring: results of a 48h fasting study with urine collection and personal activity patterns. Int J Hyg Environ Health 216, 672–681 (2013). 10.1016/j.ijheh.2012.12.002

12 Chen, L. W. et al. Modulation effects of folic acid and vitamin D on the relationships between prenatal cumulative phthalate exposure and preschoolers’ emotional and behavioral problems. Environ Int 196, 109284 (2025). 10.1016/j.envint.2025.109284

13 Cohen-Eliraz, L. et al. Prenatal exposure to phthalates and emotional/behavioral development in young children. Neurotoxicology 98, 39–47 (2023). 10.1016/j.neuro.2023.07.006

14 Engel, S. M. et al. Prenatal phthalate exposure is associated with childhood behavior and executive functioning. Environ Health Perspect 118, 565–571 (2010). 10.1289/ehp.0901470

15 England-Mason, G. et al. Similar names, different results: Consistency of the associations between prenatal exposure to phthalates and parent-ratings of behavior problems in preschool children. Environ Int 142, 105892 (2020). 10.1016/j.envint.2020.105892

16 Huang, Y. S. et al. Distinct Impacts of Prenatal and Postnatal Phthalate Exposure on Behavioral and Emotional Development in Children Aged 1.5 to 3 Years. Toxics 12 (2024). 10.3390/toxics12110795

17 Philippat, C. et al. Prenatal Exposure to Nonpersistent Endocrine Disruptors and Behavior in Boys at 3 and 5 Years. Environ Health Perspect 125, 097014 (2017). 10.1289/EHP1314

18 Tsai, T. L. et al. Co-exposure to toxic metals and phthalates in pregnant women and their children’s mental health problems aged four years - Taiwan Maternal and Infant Cohort Study (TMICS). Environ Int 173, 107804 (2023). 10.1016/j.envint.2023.107804

19 Choi, J. W. et al. Gestational phthalate exposure and behavioral problems in preschool-aged children with increased likelihood of autism spectrum disorder. Int J Hyg Environ Health 263, 114483 (2025). 10.1016/j.ijheh.2024.114483

20 Jankowska, A. et al. Prenatal and early postnatal phthalate exposure and child neurodevelopment at age of 7 years - Polish Mother and Child Cohort. Environ Res 177, 108626 (2019). 10.1016/j.envres.2019.108626

21 Minatoya, M. et al. Prenatal exposure to bisphenol A and phthalates and behavioral problems in children at preschool age: the Hokkaido Study on Environment and Children’s Health. Environ Health Prev Med 23, 43 (2018). 10.1186/s12199-018-0732-1

22 VanderWeele, T. J. Mediation Analysis: A Practitioner’s Guide. Annu Rev Public Health 37, 17–32 (2016). 10.1146/annurev-publhealth-032315-021402

23 Eisner, A. et al. Cord blood immune profile: Associations with higher prenatal plastic chemical levels. Environ Pollut 315, 120332 (2022). 10.1016/j.envpol.2022.120332

24 Frye, R. E. et al. Mitochondria May Mediate Prenatal Environmental Influences in Autism Spectrum Disorder. J Pers Med 11 (2021). 10.3390/jpm11030218

25 Thomson, S. et al. Increased maternal non-oxidative energy metabolism mediates association between prenatal di-(2-ethylhexyl) phthalate (DEHP) exposure and offspring autism spectrum disorder symptoms in early life: A birth cohort study. Environ Int 171, 107678 (2023). 10.1016/j.envint.2022.107678

26 Tanner, S. et al. A Pathway-Based Genetic Score for Oxidative Stress: An Indicator of Host Vulnerability to Phthalate-Associated Adverse Neurodevelopment. Antioxidants (Basel) 11 (2022). 10.3390/antiox11040659

27 Pham, C. et al. Maternal oxidative stress during pregnancy associated with emotional and behavioural problems in early childhood: implications for foetal programming. Mol Psychiatry 28, 3760–3768 (2023). 10.1038/s41380-023-02284-9

28 Valko, M. et al. Free radicals and antioxidants in normal physiological functions and human disease. Int J Biochem Cell Biol 39, 44–84 (2007). 10.1016/j.biocel.2006.07.001

29 Cho, Y. J., Park, S. B. & Han, M. Di-(2-ethylhexyl)-phthalate induces oxidative stress in human endometrial stromal cells in vitro. Mol Cell Endocrinol 407, 9–17 (2015). 10.1016/j.mce.2015.03.003

30 Gutierrez-Garcia, A. K., Torres-Garcia, D. A. & De Leon-Rodriguez, A. Diethyl phthalate and dibutyl phthalate disrupt sirtuins expression in the HepG2 cells. Toxicol Res (Camb) 13, tfae103 (2024). 10.1093/toxres/tfae103

31 Li, G. et al. Di(2-ethylhexyl) phthalate disturbs cholesterol metabolism through oxidative stress in rat liver. Environ Toxicol Pharmacol 95, 103958 (2022). 10.1016/j.etap.2022.103958

32 Tetz, L. M. et al. Mono-2-ethylhexyl phthalate induces oxidative stress responses in human placental cells in vitro. Toxicol Appl Pharmacol 268, 47–54 (2013). 10.1016/j.taap.2013.01.020

33 Wu, Y. et al. Oral exposure to dibutyl phthalate exacerbates chronic lymphocytic thyroiditis through oxidative stress in female Wistar rats. Sci Rep 7, 15469 (2017). 10.1038/s41598-017-15533-z

34 Zhou, D. et al. Di-n-butyl phthalate (DBP) exposure induces oxidative damage in testes of adult rats. Syst Biol Reprod Med 56, 413–419 (2010). 10.3109/19396368.2010.509902

35 Graille, M. et al. Urinary 8-OHdG as a Biomarker for Oxidative Stress: A Systematic Literature Review and Meta-Analysis. Int J Mol Sci 21 (2020). 10.3390/ijms21113743

36 Li, Y. S., Fujisawa, K. & Kawai, K. Diurnal and daily fluctuations in levels of the urinary oxidative stress marker 8-hydroxyguanosine in spot urine samples. Genes Environ 47, 1 (2025). 10.1186/s41021-025-00324-0

37 Matsumoto, Y. et al. The stability of the oxidative stress marker, urinary 8-hydroxy-2’-deoxyguanosine (8-OHdG), when stored at room temperature. J Occup Health 50, 366–372 (2008). 10.1539/joh.l7144

38 Brassea-Perez, E. et al. “Oxidative stress induced by phthalates in mammals: State of the art and potential biomarkers”. Environ Res 206, 112636 (2022). 10.1016/j.envres.2021.112636

39 Carroll, R. et al. Latent classes for chemical mixtures analyses in epidemiology: an example using phthalate and phenol exposure biomarkers in pregnant women. J Expo Sci Environ Epidemiol 30, 149–159 (2020). 10.1038/s41370-019-0181-y

40 Cathey, A. L. et al. Individual and joint effects of phthalate metabolites on biomarkers of oxidative stress among pregnant women in Puerto Rico. Environ Int 154, 106565 (2021). 10.1016/j.envint.2021.106565

41 Zeng, J. Y. et al. Prenatal exposures to phthalates and bisphenols in relation to oxidative stress: single pollutant and mixtures analyses. Environ Sci Pollut Res Int 31, 13954–13964 (2024). 10.1007/s11356-024-32032-7

42 Davalos, A. D. et al. Associations between mixtures of urinary phthalate metabolite concentrations and oxidative stress biomarkers among couples undergoing fertility treatment. Environ Res 212, 113342 (2022). 10.1016/j.envres.2022.113342

43 Huang, Y. et al. Co-exposure to organic UV filters and phthalates and their associations with oxidative stress levels in children: A prospective follow-up study in China. Sci Total Environ 905, 167433 (2023). 10.1016/j.scitotenv.2023.167433

44 Li, A. J. et al. Mediation analysis for the relationship between urinary phthalate metabolites and type 2 diabetes via oxidative stress in a population in Jeddah, Saudi Arabia. Environ Int 126, 153–161 (2019). 10.1016/j.envint.2019.01.082

45 Liu, C. et al. Mediation of the relationship between phthalate exposure and semen quality by oxidative stress among 1034 reproductive-aged Chinese men. Environ Res 179, 108778 (2019). 10.1016/j.envres.2019.108778

46 Liu, C. et al. Oxidative stress mediates the associations between phthalate exposures and thyroid cancer/benign nodule risk. Environ Pollut 326, 121462 (2023). 10.1016/j.envpol.2023.121462

47 Xiong, D. et al. Exploring the relationship between urinary phthalate metabolites and Crohn’s disease via oxidative stress, and the potential moderating role of gut microbiota: A conditional mediation model. Free Radic Biol Med 208, 468–477 (2023). 10.1016/j.freeradbiomed.2023.09.005

48 Ferguson, K. K. et al. Mediation of the Relationship between Maternal Phthalate Exposure and Preterm Birth by Oxidative Stress with Repeated Measurements across Pregnancy. Environ Health Perspect 125, 488–494 (2017). 10.1289/EHP282

49 Ortlund, K. E. et al. Oxidative stress as a potential mechanism linking gestational phthalates exposure to cognitive development in infancy. Neurotoxicol Teratol 106, 107397 (2024). 10.1016/j.ntt.2024.107397

50 Tanner, S. et al. Prenatal Environmental Determinants of Aromatase Brain-Promoter Methylation in Cord Blood: Chemical, Airborne, Pharmacological, and Nutritional Factors. bioRxiv, 2025.2007.2024.666701 (2025). 10.1101/2025.07.24.666701

51 Vuillermin, P. et al. Cohort Profile: The Barwon Infant Study. Int J Epidemiol 44, 1148–1160 (2015). 10.1093/ije/dyv026

52 Heffernan, A. L. et al. Harmonizing analytical chemistry and clinical epidemiology for human biomonitoring studies. A case-study of plastic product chemicals in urine. Chemosphere 238, 124631 (2020). 10.1016/j.chemosphere.2019.124631

53 Ponsonby, A. L. et al. Prenatal phthalate exposure, oxidative stress-related genetic vulnerability and early life neurodevelopment: A birth cohort study. Neurotoxicology 80, 20–28 (2020). 10.1016/j.neuro.2020.05.006

54 Hornung, R. W. & Reed, L. D. Estimation of average concentration in the presence of nondetectable values. Appl Occup Environ Hyg 5, 46–51 (1990).

55 Engel, S. M. et al. Prenatal Phthalates, Maternal Thyroid Function, and Risk of Attention-Deficit Hyperactivity Disorder in the Norwegian Mother and Child Cohort. Environ Health Perspect 126, 057004 (2018). 10.1289/EHP2358

56 Levine, L. & Fahy, J. P. Evaluation of urinary lead determinations. I. The significance of the specific gravity. J Ind Hyg Toxicol 27, 217–223 (1945).

57 Mortamais, M. et al. Correcting for the influence of sampling conditions on biomarkers of exposure to phenols and phthalates: a 2-step standardization method based on regression residuals. Environ Health 11, 29 (2012). 10.1186/1476-069X-11-29

58 Achenbach, T. M. & Rescorla, L. Manual for the ASEBA preschool forms & profiles. (University of Vermont Research Center for Children, Youth and Families, 2000).

59 Achenbach, T. M., Dumenci, L. & Rescorla, L. A. DSM-oriented and empirically based approaches to constructing scales from the same item pools. J Clin Child Adolesc Psychol 32, 328–340 (2003). 10.1207/S15374424JCCP3203_02

60 Lacalle Sistere, M., Domenech Massons, J. M., Granero Perez, R. & Ezpeleta Ascaso, L. Validity of the DSM-Oriented scales of the Child Behavior Checklist and Youth Self-Report. Psicothema 26, 364–371 (2014). 10.7334/psicothema2013.342

61 Goodman, R. The Strengths and Difficulties Questionnaire: a research note. J Child Psychol Psychiatry 38, 581–586 (1997). 10.1111/j.1469-7610.1997.tb01545.x

62 Stone, L. L., Otten, R., Engels, R. C., Vermulst, A. A. & Janssens, J. M. Psychometric properties of the parent and teacher versions of the strengths and difficulties questionnaire for 4- to 12-year-olds: a review. Clin Child Fam Psychol Rev 13, 254–274 (2010). 10.1007/s10567-010-0071-2

63 Armitage, J. M. et al. Validation of the Strengths and Difficulties Questionnaire (SDQ) emotional subscale in assessing depression and anxiety across development. PLoS One 18, e0288882 (2023). 10.1371/journal.pone.0288882

64 Carrico, C., Gennings, C., Wheeler, D. C. & Factor-Litvak, P. Characterization of Weighted Quantile Sum Regression for Highly Correlated Data in a Risk Analysis Setting. J Agric Biol Environ Stat 20, 100–120 (2015). 10.1007/s13253-014-0180-3

65 Tanner, E. M., Bornehag, C. G. & Gennings, C. Repeated holdout validation for weighted quantile sum regression. MethodsX 6, 2855–2860 (2019). 10.1016/j.mex.2019.11.008

66 Busgang, S. A. et al. Application of growth modeling to assess the impact of hospital-based phthalate exposure on preterm infant growth parameters during the neonatal intensive care unit hospitalization. Sci Total Environ 850, 157830 (2022). 10.1016/j.scitotenv.2022.157830

67 gWQS: Generalized Weighted Quantile Sum Regression v. R package version 3.0.5 (2023).

68 Steen, J., Loeys, T., Moerkerke, B. & Vansteelandt, S. medflex: An R Package for Flexible Mediation Analysis using Natural Effect Models. Journal of Statistical Software 76, 1–46 (2017). 10.18637/jss.v076.i11

69 Bobb, J. F. et al. Bayesian kernel machine regression for estimating the health effects of multi-pollutant mixtures. Biostatistics 16, 493–508 (2015). 10.1093/biostatistics/kxu058

70 bkmr: Bayesian Kernel Machine Regression v. R package version 0.2.2 (2022).

71 bkmrhat: Parallel Chain Tools for Bayesian Kernel Machine Regression v. R package version 1.1.3 (2022).

72 Plummer, M., Best, N., Cowles, K. & Vines, K. CODA: Convergence Diagnosis and Output Analysis for MCMC. R News 6, 7–11 (2006).

73 van Buuren, S. & Groothuis-Oudshoorn, K. mice: Multivariate Imputation by Chained Equations in R. Journal of Statistical Software 45, 1–67 10.18637/jss.v045.i03

74 White, I. R., Royston, P. & Wood, A. M. Multiple imputation using chained equations: Issues and guidance for practice. Stat Med 30, 377–399 (2011). 10.1002/sim.4067

75 Dawson, S. L. et al. Maternal prenatal gut microbiota composition predicts child behaviour. EBioMedicine 68, 103400 (2021). 10.1016/j.ebiom.2021.103400

76 Reddam, A., McLarnan, S. & Kupsco, A. Environmental Chemical Exposures and Mitochondrial Dysfunction: a Review of Recent Literature. Curr Environ Health Rep 9, 631–649 (2022). 10.1007/s40572-022-00371-7

77 Yeldandi, A. V., Rao, M. S. & Reddy, J. K. Hydrogen peroxide generation in peroxisome proliferator-induced oncogenesis. Mutat Res 448, 159–177 (2000). 10.1016/s0027-5107(99)00234-1

78 Choi, K. et al. In vitro metabolism of di(2-ethylhexyl) phthalate (DEHP) by various tissues and cytochrome P450s of human and rat. Toxicol In Vitro 26, 315–322 (2012). 10.1016/j.tiv.2011.12.002

79 Veith, A. & Moorthy, B. Role of Cytochrome P450s in the Generation and Metabolism of Reactive Oxygen Species. Curr Opin Toxicol 7, 44–51 (2018). 10.1016/j.cotox.2017.10.003

80 Jiao, Y. et al. Monobutyl phthalate (MBP) can dysregulate the antioxidant system and induce apoptosis of zebrafish liver. Environ Pollut 257, 113517 (2020). 10.1016/j.envpol.2019.113517

81 Mohammadi, H. & Ashari, S. Mechanistic insight into toxicity of phthalates, the involved receptors, and the role of Nrf2, NF-kappaB, and PI3K/AKT signaling pathways. Environ Sci Pollut Res Int 28, 35488–35527 (2021). 10.1007/s11356-021-14466-5

82 Ferguson, K. K. et al. Repeated measures of urinary oxidative stress biomarkers during pregnancy and preterm birth. Am J Obstet Gynecol 212, 208 e201-208 (2015). 10.1016/j.ajog.2014.08.007

83 Holland, N. et al. Urinary Phthalate Metabolites and Biomarkers of Oxidative Stress in a Mexican-American Cohort: Variability in Early and Late Pregnancy. Toxics 4 (2016). 10.3390/toxics4010007

84 Waits, A. et al. Urinary phthalate metabolites are associated with biomarkers of DNA damage and lipid peroxidation in pregnant women - Tainan Birth Cohort Study (TBCS). Environ Res 188, 109863 (2020). 10.1016/j.envres.2020.109863

85 Zhang, Y. J. et al. DNA oxidative damage in pregnant women upon exposure to conventional and alternative phthalates. Environ Int 156, 106743 (2021). 10.1016/j.envint.2021.106743

86 Rommel, A. S. et al. Associations between urinary biomarkers of oxidative stress in the third trimester of pregnancy and behavioral outcomes in the child at 4 years of age. Brain Behav Immun 90, 272–278 (2020). 10.1016/j.bbi.2020.08.029

87 Peter Stein, T., et al. Oxidative stress early in pregnancy and pregnancy outcome. Free Radic Res 42, 841–848 (2008). 10.1080/10715760802510069

88 Buss, C. Maternal oxidative stress during pregnancy and offspring neurodevelopment. Brain Behav Immun 93, 6–7 (2021). 10.1016/j.bbi.2021.01.007

89 Murphy, B. E., Clark, S. J., Donald, I. R., Pinsky, M. & Vedady, D. Conversion of maternal cortisol to cortisone during placental transfer to the human fetus. Am J Obstet Gynecol 118, 538–541 (1974). 10.1016/s0002-9378(16)33697-3

90 Zaretsky, M. V., Alexander, J. M., Byrd, W. & Bawdon, R. E. Transfer of inflammatory cytokines across the placenta. Obstet Gynecol 103, 546–550 (2004). 10.1097/01.AOG.0000114980.40445.83

91 Woods, R. M. et al. Maternal immune activation and role of placenta in the prenatal programming of neurodevelopmental disorders. Neuronal Signal 7, NS20220064 (2023). 10.1042/NS20220064

92 Chambers, J. C. et al. Association of maternal endothelial dysfunction with preeclampsia. JAMA 285, 1607–1612 (2001). 10.1001/jama.285.12.1607

93 Henderson, G. I., Chen, J. J. & Schenker, S. Ethanol, oxidative stress, reactive aldehydes, and the fetus. Front Biosci 4, D541–550 (1999). 10.2741/henderson

94 Buss, C. et al. Maternal cortisol over the course of pregnancy and subsequent child amygdala and hippocampus volumes and affective problems. Proc Natl Acad Sci U S A 109, E1312–1319 (2012). 10.1073/pnas.1201295109

95 Guilbert, A. et al. Associations between a mixture of phenols and phthalates and child behaviour in a French mother-child cohort with repeated assessment of exposure. Environ Int 156, 106697 (2021). 10.1016/j.envint.2021.106697

96 Dewey, D. et al. Sex-specific associations between maternal phthalate exposure and neurodevelopmental outcomes in children at 2 years of age in the APrON cohort. Neurotoxicology 98, 48–60 (2023). 10.1016/j.neuro.2023.07.005

97 Khalfallah, O. et al. Cytokines as mediators of the associations of prenatal exposure to phenols, parabens, and phthalates with internalizing behaviours at age 3 in boys: A mixture exposure and mediation approach. Environ Res 229, 115865 (2023). 10.1016/j.envres.2023.115865

98 European Parliament. Regulation (EC) No 1907/2006 of the European Parliament and of the Council of 18 December 2006 concerning the Registration, Evaluation, Authorisation and Restriction of Chemicals (REACH). (2006). <https://eur-lex.europa.eu/eli/reg/2006/1907/oj/eng>

99 United States Government. The Consumer Product Safety Improvement Act of 2008. Public Law 110–314. (2008). <https://www.cpsc.gov/s3fs-public/cpsia.pdf>

100 European Parliament. Regulation (EC) No 1223/2009 of the European Parliament and of the Council of 30 November 2009 on cosmetic products. (2009). <https://eur-lex.europa.eu/legal-content/EN/TXT/?uri=CELEX%3A32009R1223&qid=1757137345964>

101 Government of Canada. Phthalates Regulations. SOR/2016-188. (2016). <https://laws-lois.justice.gc.ca/PDF/SOR-2016-188.pdf>

102 Australian Government. Therapeutic Goods (Poisons Standard—June 2025) Instrument 2025. (2025). <https://www.legislation.gov.au/F2025L00599/asmade/text>

103 Eichler, C. M. A., Cohen Hubal, E. A. & Little, J. C. Assessing Human Exposure to Chemicals in Materials, Products and Articles: The International Risk Management Landscape for Phthalates. Environ Sci Technol 53, 13583–13597 (2019). 10.1021/acs.est.9b03794

104. European Chemicals Agency (ECHA). Assessment of regulatory needs: Ortho-phthalates. (2021). <https://echa.europa.eu/documents/10162/f0b44d3c-b9fa-b3af-f211-b3feb8055ac3>

105 Adibi, J. J. et al. Characterization of phthalate exposure among pregnant women assessed by repeat air and urine samples. Environ Health Perspect 116, 467–473 (2008). 10.1289/ehp.10749

106 Suzuki, Y. et al. Exposure assessment of phthalate esters in Japanese pregnant women by using urinary metabolite analysis. Environ Health Prev Med 14, 180–187 (2009). 10.1007/s12199-009-0078-9

107 Li, Y. S. et al. Diurnal and day-to-day variation of urinary oxidative stress marker 8-hydroxy-2’-deoxyguanosine. J Clin Biochem Nutr 68, 18–22 (2021). 10.3164/jcbn.19-105

108 Sule, R. O., Rivera, G. D. T., Vaidya, T., Gartrell, E. & Gomes, A. V. Environmental Toxins and Oxidative Stress: The Link to Cardiovascular Diseases. Antioxidants (Basel) 14 (2025). 10.3390/antiox14050604

