## Supplementary figures for "Prenatal phthalate mixture exposure increases early childhood internalising problems via maternal oxidative stress"

*Table of contents*

**Figure S1.** Barwon Infant Study participant flowchart

**Figure S2.** Distributions of mean WQS regression and mediation estimates (averaged across holdouts) across the imputed datasets

**Figure S3.** Distribution of mean WQS component weight estimates (averaged across holdouts) across the imputed datasets

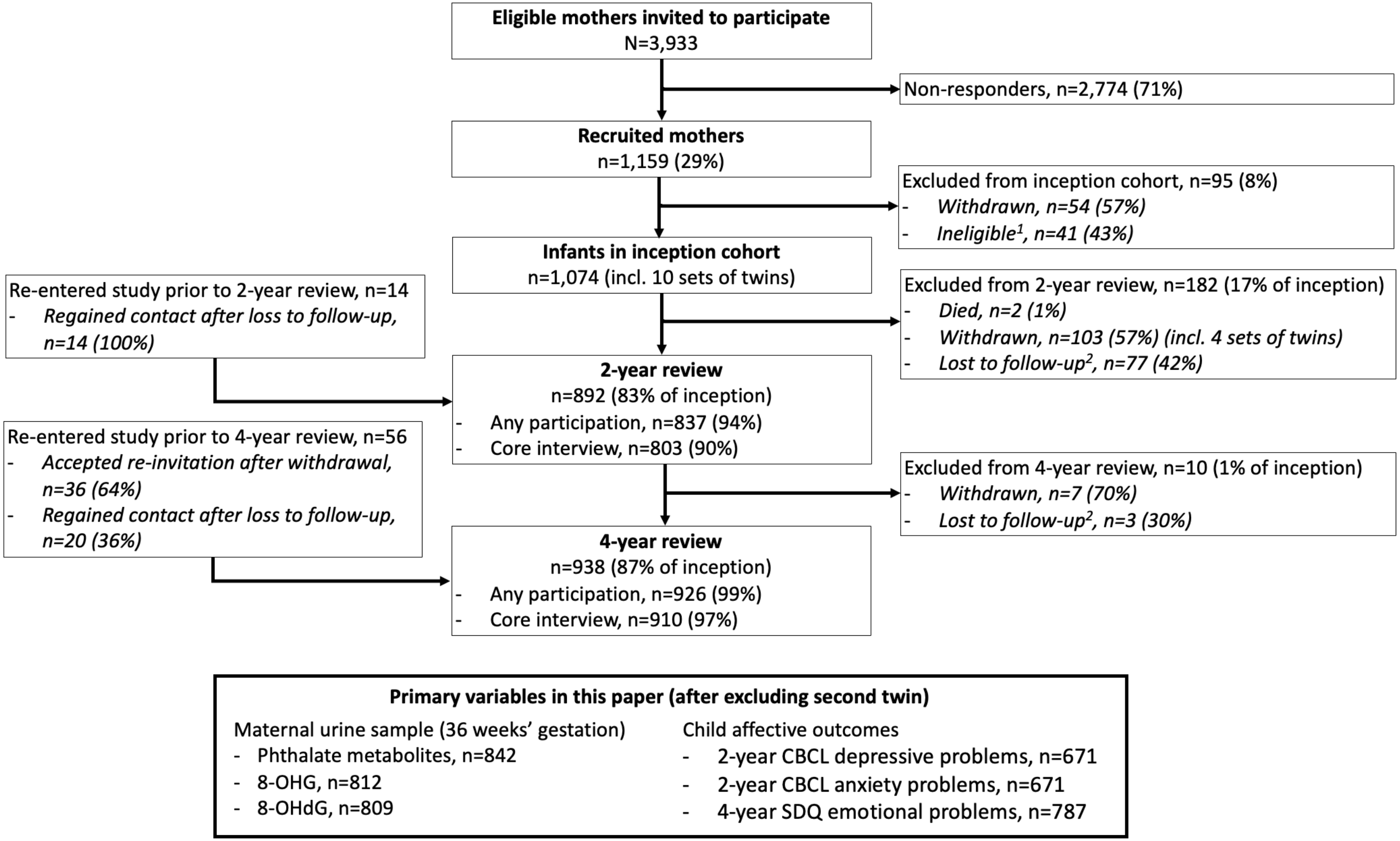


**Figure S1.** Barwon Infant Study participant flowchart

[1] Due to: no longer being resident in Barwon region (n=12), <32 weeks' gestation (n=8), serious illness in first few days of life (n=7), major congenital disease (n=5), stillbirth (n=5), miscarriage (n=2), or cord blood stored privately (n=2)

[2] Loss to follow-up defined as missing two consecutive reviews

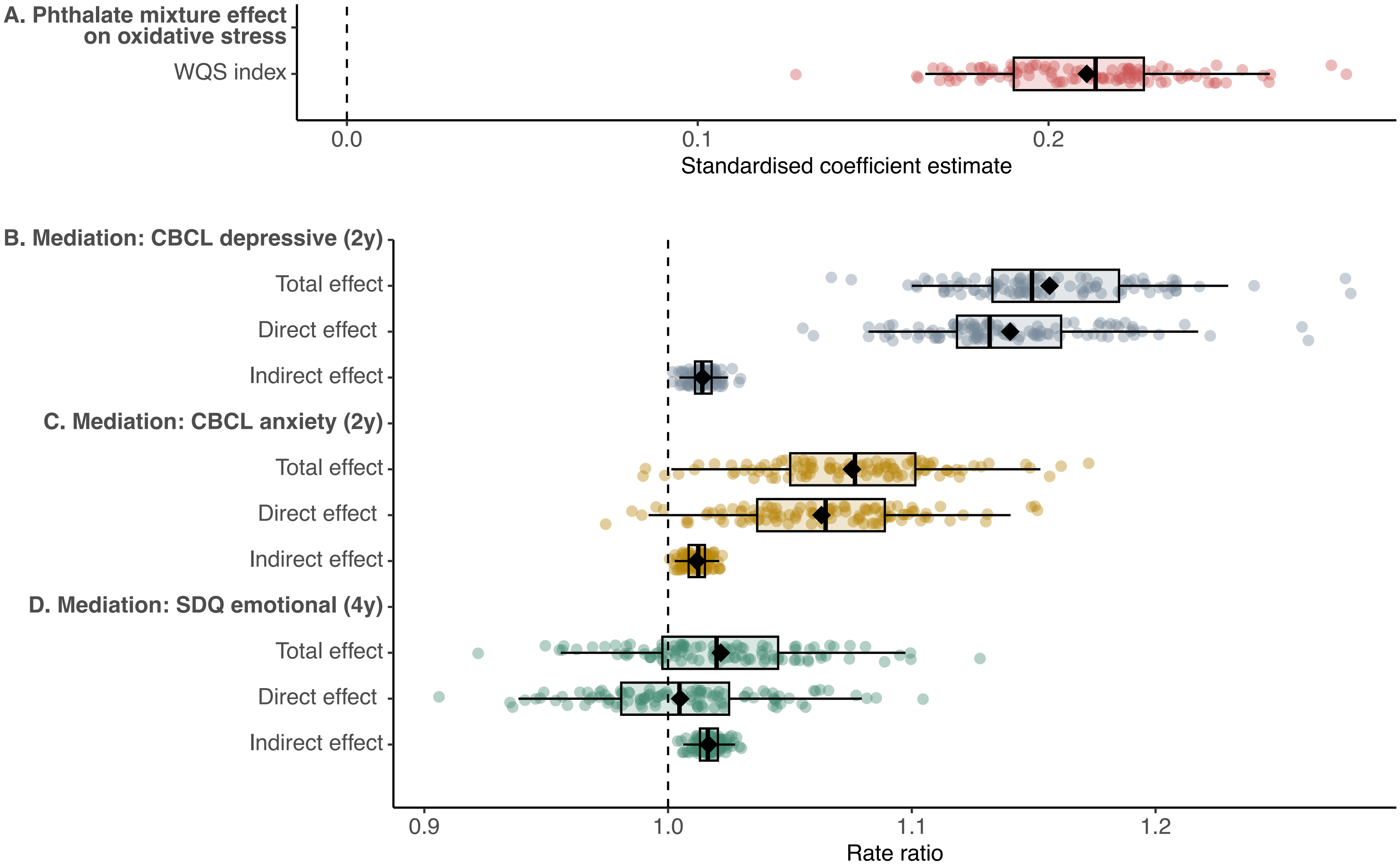


**Figure S2.** Estimated phthalate mixture effect on maternal oxidative stress in pregnancy from weighted quantile sum regression analysis with repeated holdouts in 100 multiply imputed datasets (**A**). Causal mediation analysis estimates in 100 holdouts in each of 100 multiply imputed datasets, with corresponding WQS index from analysis A as exposure, oxidative stress as mediator, and child CBCL DSM-5-oriented depressive problems subscale score at 2 years as outcome (**B**) or child CBCL DSM-5-oriented anxiety problems subscale score at 2 years as outcome (**C**) or child SDQ emotional problems subscale score at 4 years as outcome (**D**). Each data point is the standardised mean estimate across the 100 holdouts for an imputed dataset. Box plots summarise the distribution of the mean estimates across the 100 imputed datasets (box represents the 25th, 50th, and 75th percentiles, and whiskers extend to the 2.5th and 97.5th percentiles). Closed diamonds represent the overall mean estimates.

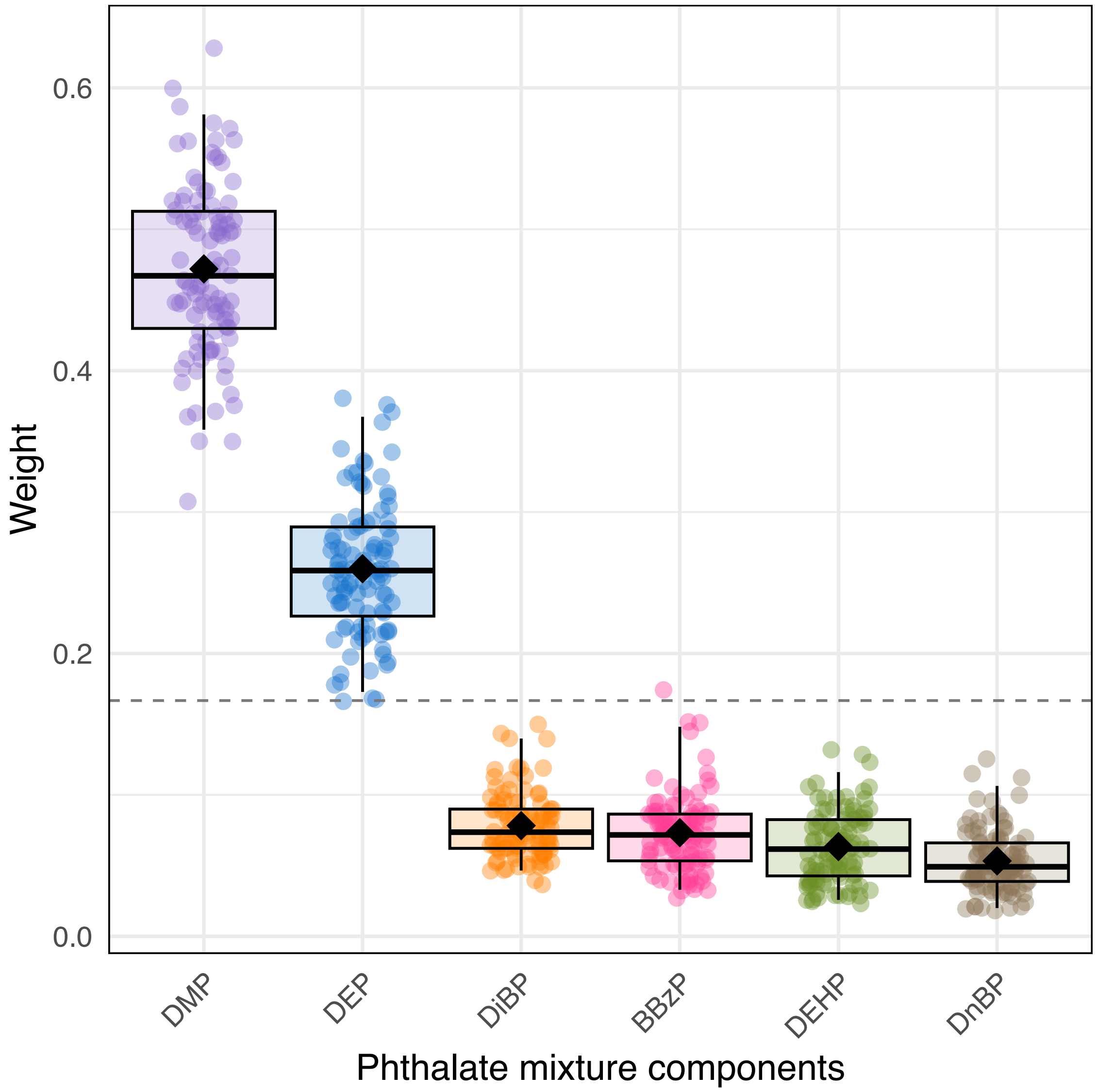


**Figure S3.** Identification of phthalate chemicals of concern in relation to maternal oxidative stress in pregnancy using weighted quantile sum regression with repeated holdouts in multiply imputed data. Each data point represents the mean weight, averaged across 100 holdouts, in one of 100 imputed datasets. Box plots summarise the distribution of the mean weights across the 100 imputed datasets (box represents the 25th, 50th, and 75th percentiles, and whiskers extend to the 2.5th and 97.5th percentiles). Closed diamonds represent the mean of the mean weights. The dotted line shows the expected weight (1/6=0.17) if all six phthalate compounds contributed equally to the mixture. *Abbreviations:* DMP, dimethyl phthalate; DEP, diethyl phthalate; DiBP, diisobutyl phthalate; BBzP, butyl benzyl phthalate; DEHP, di-(2-ethylhexyl) phthalate; DnBP, di-n-butyl phthalate.
